# Prevalence of Hepatitis B, Hepatitis C and HIV infection among patients undergoing hemodialysis in Argentina

**DOI:** 10.1101/2020.08.04.20168385

**Authors:** Matías J. Pereson, Alfredo P. Martínez, Katia Isaac, Gustavo Laham, Ezequiel Ridruejo, Gabriel H. Garcia, Diego M. Flichman, Federico A. Di Lello

## Abstract

**Background and Aim:** Blood-borne infections are a major cause of damage in individuals on hemodialysis (HD). In particular, knowledge about the viral status of hepatitis B (HBV), hepatitis C (HCV) and human immunodeficiency virus (HIV) in HD patients is gold data to face medical challenges. Related information is scarce in Argentina. The aim of this study was to determine the prevalence of HBV, HCV and HIV infections in HD patients from Buenos Aires, Argentina.

**Methods:** Seven hundred and forty-eight HD patients were included in a retrospective cross-sectional study. Laboratories assays were performed to determine virological status. HCV genotyping was carried out by phylogenetic analysis of NS5B partial gene.

**Results:** Infection with one of the viruses was detected in 31.1% of patients [HBV in 82 (11.0%), HCV in 179 (23.9%), and HIV in 6 (0.8%)]. Thirty-two (4.3%) patients had two virus markers [27 (3.6%) with HCV/HBV, 4 (0.5%) with HCV/HIV and 1 (0.13%) with HBV/HIV]. Finally, one single patient (0.13%) presented all three markers. Time on dialysis was correlated with HCV infection but no with HBV. Distribution of HCV subtypes was inverted respect to the observed in general population [HCV-1a (73.2%) and HCV-1b (26.8%) in HD vs. HCV-1a (26.5%) and HCV-1b (73.5%) in general population, p<0.001].

**Conclusions:** These infections, mainly HCV, continue to occur at a very high rate in HD patients. Results emphasize the need to consider a priority the HCV infected patients in dialysis treatment and to vaccine against HBV in order to decrease its morbidity and mortality.

## Introduction

The prevalence rate of dialyzed patients in Argentina is 637 patients per million individuals (approximately 29,700 patients) being hemodialysis (HD) the most widely applied treatment for patients with end-stage renal disease [1]. Blood-borne viruses, such as hepatitis B (HBV), hepatitis C (HCV) and human immunodeficiency (HIV), have a consistently higher prevalence among HD patients than in general population [2]. This finding may be due to several factors including the repeated and prolonged access to the bloodstream, the simultaneous treatment of multiple patients in the same facility and the lack of effective biosafety measures. This issue is a major healthcare concern since these viruses are a significant cause of increased morbidity and mortality [3]. Prevalence rate varies among countries and even among health centers within a same area [2, 4-6]. Coinfection is a common finding among HD patients and it may be associated to worst clinical evolutions [7-9].

There are not large-scale general population prevalence studies for HBV, HCV, and HIV in Argentina. Flichman et al. (2014), informed a low infection prevalence in blood donors tested nationwide [10]. This authors analyzed, in more than two million samples, prevalence for HBsAg (0.19%), anti-HCV (0.46%), and anti-HIV (0.20%) titers.

As regards the Argentine population undergoing dialysis, data is unfortunately fragmented and dissimilar. The Argentine Chronic Dialysis Registry reports prevalence of 1.02%, 2.40%, and 0.91% for HBsAg, anti-HCV, and anti-HIV, respectively [1]. Other studies conducted on different dialysis populations from Argentina showed an HCV significantly higher prevalence, ranging from 10.6 to 38.0% [11-14]. Disparities in reported HCV prevalence rates as well as paucity of data for HBV and HIV warrants the present research work. Therefore, the main goal of this study was to estimate the prevalence of HBV, HCV, and HIV among HD patients from the Ciudad Autónoma de Buenos Aires, Argentina.

## Materials and Methods

### Study population

In this retrospective cross-sectional study, patients in hemodialysis were tested for anti-HCV, HBsAg, and anti-HIV as part of a screening program conducted at Centro de Educación Médica e Investigaciones Clínicas Norberto Quirno (CEMIC), a university hospital located in Buenos Aires, Argentina. Samples were collected between January 2001 and December 2018.

### Laboratory assays

HBV serological markers were analyzed with AxSYM, Abbott Diagnostics, USA (samples before 2010) and with Architect Abbott system, Abbott Diagnostics, Wiesbaden, Germany (samples since 2010). Anti-HCV testing was performed by anti-HCV enzyme immunoassay AxSYM HCV V3.0 (Abbott, Wiesbaden, Germany) and ARCHITECT anti-HCV (Abbott Wiesbaden, Germany). HIV was tested by AxSYM HIV 1/2 gO (Abbott, Wiesbaden, Germany) and ARCHITECT HIV Ag/Ab Combination (Abbott Wiesbaden, Germany).

### HCV genotyping, RT-PCR and sequencing

Serum RNAs were extracted with Magna Pure (Roche) following the manufacturer‘s instructions. Reverse transcription was carried out with MMLV-RT (Promega) with Random Hexamer Primers using the manufacturer‘s protocol. The amplification of HCV NS5B partial region was performed as previously described [15]. Subsequently, the amplified DNAs were purified and then sequenced in both senses (Instituto Nacional de Tecnología Agropecuaria, Castelar, Argentina).

### Phylogenetic analysis and HCV genotype

A broad sequence search was performed using keywords on the NCBI central nucleotide search website (https://blast.ncbi.nlm.nih.gov/Blast.cgi?PROGRAM=blastn&PAGE_TYPE=BlastSearch&LINK_LOC=blasthome) to retrieve sequences of HCV containing NS5B regions. One hundred fifty nine sequences were randomly selected so that all HCV genotypes and each of the geographic regions of interest were represented. NS5B sequences were aligned with CLUSTALX v2.0 software. A Maximum-Likelihood (ML) phylogenetic tree was estimated in MEGA-X setting the model parameters according to those suggested by Bayesian Information Criterion [16]. A non-parametric bootstrap analysis of 1000 replicates was carried out for branch support. In order to verify the obtained results, a Bayesian phylogenetic analysis was simultaneously carried out with MrBayes v3.2.7a [17]. Analyses were run for five million generations, and sampled every 5000 generations. Convergence of parameters (effective sample size (ESS) ≥ 200, with a 10% burn-in) was verified with Tracer v1.7.1. Phylogenetic trees were visualized with FigTree v1.4.4.

### Statistical analysis

Frequencies were compared using the chi-square test or the Fisher‘s test. The Student‘s t-test and the Mann-Whitney U were used for comparing continuous variables. The statistical analysis was carried out using the SPSS statistical software package release 19.0 (IBM SPSS Inc., Chicago, IL, USA).

### Nucleotide sequences accession numbers

Nucleotide sequences for the HCV have been deposited in GenBank under accession numbers MK187268/272/278/287/292-294/298/299/301/330/338 and MN982201-MN982231.

### Ethical aspects

Written informed consents to participate in the study were obtained from patients. The study protocol was approved by the ethics committee from “Facultad de Farmacia y Bioquímica, Universidad de Buenos Aires” (record number 02032015-2/2015) in accordance with the 1975 Helsinki Declaration.

## Results

### Characteristics of the study population

A cohort of 748 patients were analyzed, median (Q1-Q3) age was 59 (44-70) years old and 410 (54.8%) were male. Sixty-three out of 748 (8.4%) were kidney transplanted patients and 182 (24.3%) had received blood transfusion. Median (Q1-Q3) time on dialysis treatment was 25 (7-76) months.

### Prevalence of HBV, HCV, and HIV infection

Infection with one of the three viruses was detected in 233 (31.1%) subjects from the cohort. Thus, 515 (68.9) patients were not infected by any of the studied viruses. The median age (Q1-Q3) of uninfected patients was 61 (48-73) and 292 (56.7) were male. Table 1 summarizes the characteristics of the study population according to the infection by any of the three considered viruses. Thirty-two (4.27%) patients had markers of infection with two virus types [27 (3.6%) with HCV/HBV, 4 (0.5%) with HCV/HIV and 1 (0.13%) with HBV/HIV]. Finally, only one patient (0.13%) presented all three virus types markers.

**Table 1.**
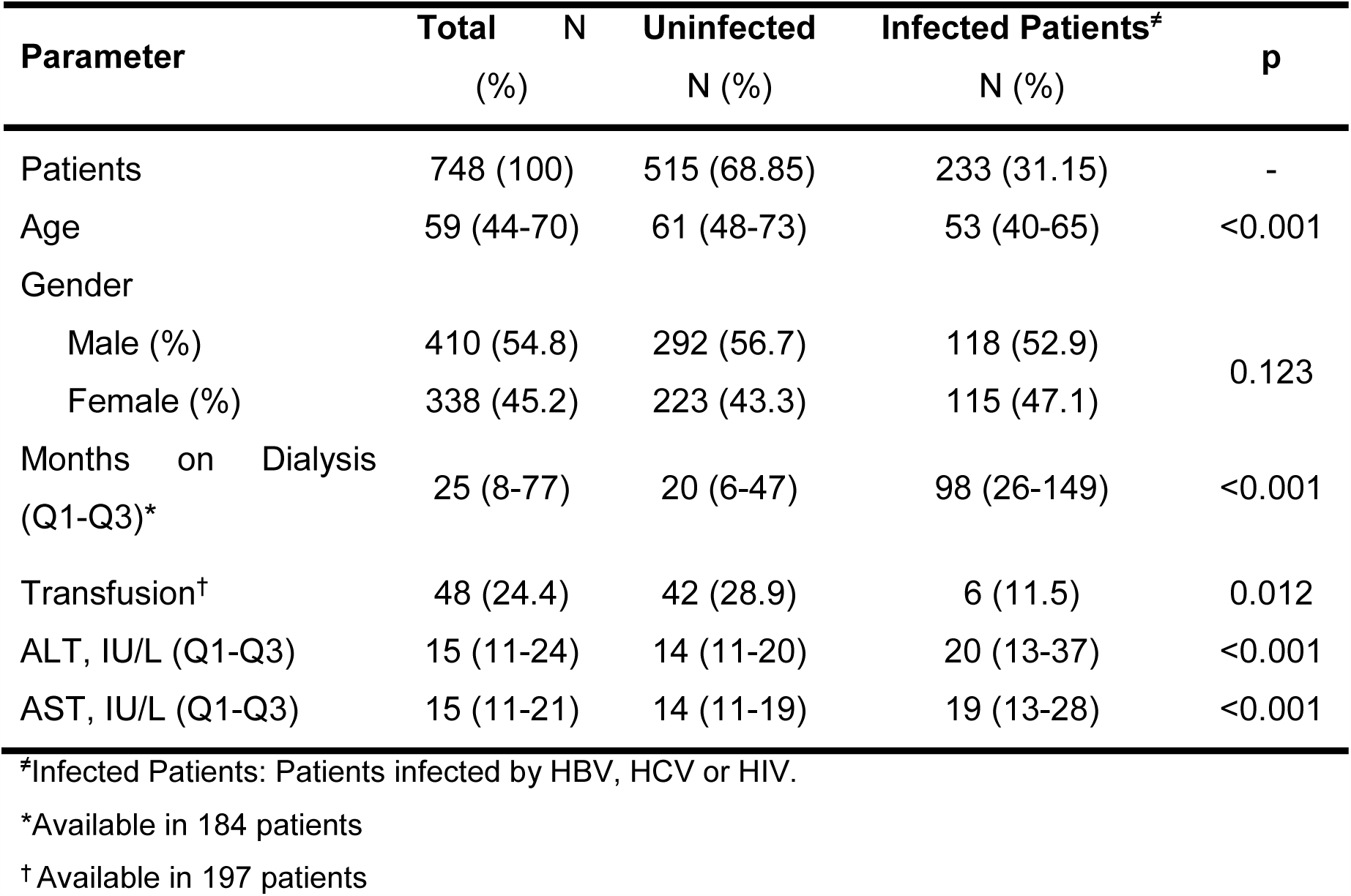
Characteristic of the study population by infection.

The HBsAg was detected in 82 patients (11.0%), 43 female and 39 male. Thus there was a similar distribution of positive cases between genders [12.7% female vs. 9.5% male, (p = 0.162)]. Neither the patient age nor the time on dialysis was statistically associated with HBV infection. In this sense, patients infected with HBV were 60 (44-69) years old and the uninfected ones were 58 (44-71) years old, p = 0.984. When considering the time on dialysis, patients infected with HBV had been on dialysis for 19 (4-156) months while those uninfected had attended dialysis for 26 (8-74) months, p = 0.927. No association was found between transfusion history and the risk of acquiring HBV [2 transfused individuals (4.2%) and 10 non-transfused ones (6.7%), (p = 0.521)]. There was no association between HBV detection and plasma levels of alanine aminotransferase (ALT) and aspartate aminotransferase (AST). Thus, subjects infected with HBV showed ALT 15 IU/L (11-26) and AST 17 IU/L (11-23), whereas the corresponding figures among non-HBV infected patients were 15 IU/L (11-24) for ALT, and 15 IU/L (11-21) for AST (p=0.911 and p=0.572 for ALT and AST, respectively).

As regards HCV, anti-HCV was detected in 179 (23.9%) patients. Plasma HCV-RNA, indicative of active HCV infection, was detected in 125 (69.8%) patients. There was no significant gender related-variation for positive cases [80 females (23.7%) and 99 males (24.1%)] (p = 0.879). Unlike findings for HBV, HCV infection in dialyzed patients was statistically associated with age [62 (48-72) years old for the uninfected subjects and 49 (39-61) years old for the infected ones, p <0.001], and with the time on dialysis [19 (6-46) months for uninfected patients and 101 (31-154) months for the infected ones, p <0.001]. In addition, subjects without transfusion history concentrated the highest number of HCV cases [40 non-transfused individuals (26.8%) versus 5 transfused (10.4%), p = 0.018]. Finally, ALT and AST were higher in patients infected with HCV. Thus, patients with HCV showed ALT 24 IU/L (15-41) and AST 21 IU/L (14-32), whereas levels for non-HCV infected patients were 14 IU/L (11-20) for ALT, and 14 IU/L (11-19) for AST (p<0.001 for both ALT and AST).

HIV presented the lowest prevalence. Only 6 (0.8%) patients were positive for anti-HIV. Statistical analysis for age, years of attending dialysis, gender, or transfusion history was not possible due to the small number of positive cases.

### HCV genotype distribution and phylogenetic analysis

As result of the particularly high prevalence of HCV infection found in this work, a phylogenetic analysis of HCV sequences was carried out in order to determine and compare the genotype of HCV infecting dialyzed patients, to those infected individuals from general population.

Nucleotide sequences from NS5B region of 43 out of 179 HCV positive cases were analyzed. The genotype distribution obtained by phylogenetic analysis was 30 (73.2%) HCV-1a, 11 (26.8) HCV-1b, 1 (2.3%) HCV-2c, and 1 (2.3%) HCV-3a.

NS5B sequences from dialyzed patients infected with HCV-1 were compared with 68 HCV-1 infected patients who attended the same hospital but not undergo HD during the same period. In this cohort, the distribution of subtypes was significantly different: 50 (73.5%) HCV-1b and 18 (26.5%) HCV-1a, p<0.001.

The patient age was statistically associated with HCV-1 subtypes distribution [51 (47-58) years old for HCV-1a 59 (50-66) years old for the infected with HCV-1b, p=0.008]. On the other hand, there was no significant differences in age for HD patients and patients attended the same hospital [51 (45-64) years old and 57 (49-62) years old, respectively, p = 0.293].

Phylogenetic analysis did not show the formation of supported clusters belonging to the same subtype between the sequences of patients undergoing dialysis and those not dialyzed (Figure 1).

**Figure 1:**
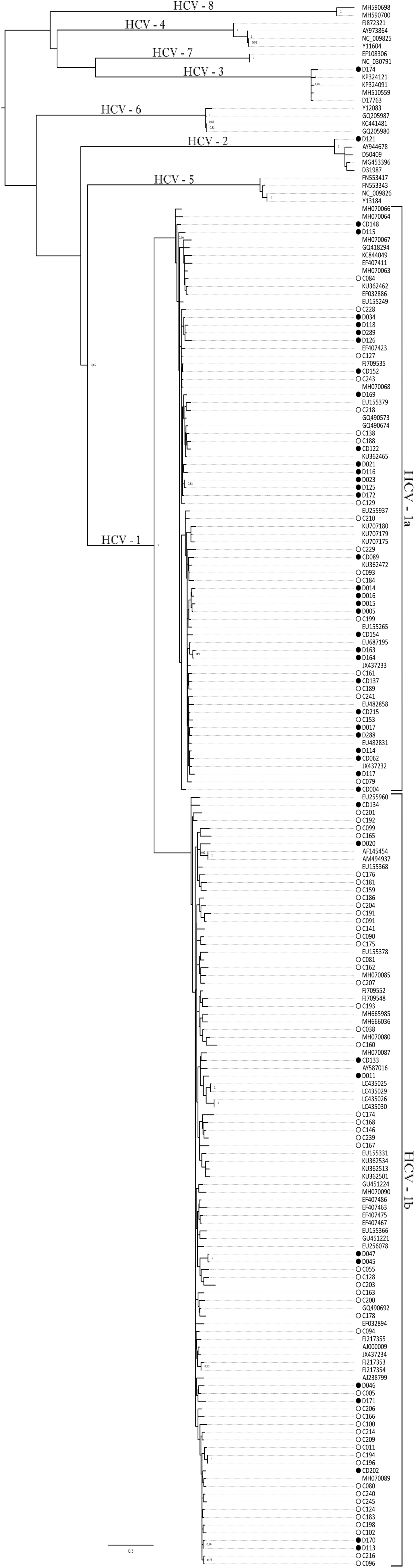
HCV maximum likelihood phylogenetic tree (T92+I+G model for nucleotide substitutions) of NS5B partial region (314nt). Branch lengths are proportional to the evolutionary distance between the samples. Numbers on each node represent the bootstrap proportion support (values lower than 0.7 are not shown). Samples were identified as follows: sequences obtained on this work have symbols before their names that represent the source: solid circle, dialysis; empty circle, no dialysis; sequences without symbols are those randomly chosen from Genbank.

## Discussion

The determination of blood-borne infections prevalence in the hemodialysis setting is relevant to establish the status of the population undergoing dialysis and to monitor the effectiveness of implemented sanitary and strategies as well as to identifying their achievements and limitations. To our knowledge, this is the first large scale survey that analyzes HBV, HCV, and HIV viral markers in a population undergoing hemodialysis in Argentina.

It is well known that dialysis procedure plays a preponderant role in the transmission of HBV and that prevalence mostly depends on the baseline population rates [2, 18]. In developed countries, HBV prevalence in dialysis centers varies between 0 - 6.6% while in developing countries it widely ranges between 1.3 and 14.6% [18]. In the present study, the HBsAg prevalence was consistent with that found in low-income countries. This high prevalence could be a consequence of the low coverage rate of the HBV vaccine (less than 40%) in the population not reached by mandatory vaccination [1]. Other reason may be an upward trend of new HBV cases, observed in the last 10 years, in people 20 to 59 years old, which represented a large proportion (49%) of individuals infected with HBV in this study [19]. In this scenario, it is crucial to increase the HBV vaccination coverage and to improve the standard precautions and/or dialysis specific procedures in order to prevent disease dissemination in our country.

Interestingly, no association was found between HBV infection and the time on dialysis. A possible explanation may be given by the fact that, as it is known in our country; most patients who start dialysis are not vaccinated against HBV. However, once they start treatment, they are immediately vaccinated. In this way, it has been observed that protection levels against HBV are approximately 30% in the first 5 months of dialysis and reach up to 80% at 15 years under treatment [1].

With regard to HCV, almost a quarter of analyzed patients were positive. This prevalence is within the previously reported range in our country for patients undergoing dialysis and significantly higher than that described in general population [13, 14, 20, 21]. This situation repeats all over the world, being people who attend HD centers a large risk group of acquiring HCV [2]. In addition, prevalence was associated with time on dialysis. In this sense, the majority of HCV-infected patients were younger but with a longer treatment history. This data is coincident with that previously published also reporting that dialyzed patients do not have the same age epidemiological distribution for HCV infections. A similar situation has been previously described in Argentina [1, 14] and other world regions [2].

HCV genotype distribution in HD patients is a valuable tool to understand the transmission routes. Some studies have found a matching distribution of genotypes among patients undergoing dialysis and general population, while others observed a frequency bias [22]. In Argentina, HCV-1b has been reported as the most prevalent subtype, both in general population and in dialyzed patients [12, 23]. This subtype has been associated with older individuals and patients who received transfusions [24]. Unexpectedly, in the cohort included in this study, a high prevalence of HCV-1a, which has been mostly found in younger individuals and intravenous drug users, was observed [24, 25]. However, in the present study, even when HD patients were younger, no significant differences were found between both populations (HD and ambulatory patients). Therefore, this finding could be a consequence of the epidemiological change of HCV in Argentina, where it has been described that although HCV-1b entered earlier in the country, it is currently on a demographic plateau, while HCV-1a was introduced later but is still in a growth phase [26]. Additionally, in patients on dialysis, a lower prevalence of HCV-2 and HCV-3 have been detected, compared to those previously reported in the general population [14, 27].

As stated for HBV, the prevalence of HIV in patients undergoing dialysis largely depends on the local prevalence in general population^4^. In this study, the observed prevalence has been higher than that described in general population but close to that reported by the Argentine Chronic Dialysis Registry [1, 28].

In Argentina, nosocomial outbreaks of HIV have not been described since 1993 [11]. As a result, the positive cases found in this study may be a consequence of the progression to end stage renal disease (ERSD) due to HIV infection rather than a consequence of the dialysis procedure itself. It is well known that risk of progression to ERSD is significantly higher in seropositive patients [4, 29]. The progression to ERSD has increased in recent years due to the higher longevity of HIV infected patients, thanks to antiretroviral therapy efficacy [30].

This work has some limitations. One of them is the lack of information about of HBV, HCV, and HIV patient status prior to dialysis. However, when considering data in general population, we can attribute the identified high prevalence to risk associated to HD. In second place, time undergoing treatment is unknown for some patients. Still, we had sufficient data to perform the statistical analysis. Lastly, samples included in this study came from a single center and prevalence can vary significantly between dialysis units [2, 4-6]. Therefore, despite the size of the cohort, caution should be taken to extrapolate results to the general problem of infections in this population.

Overall, almost one third of the studied patients presented one of the three analyzed viruses. Unfortunately, the implementation of universal precautionary biosafety standards on dialysis units would appear to be insufficient to contain blood-borne infections. Further efforts, such as HBV vaccination and HCV treatment with new drugs, will be required to prevent HD patients from continuing to be a vulnerable population at high risk of acquiring blood-borne infections. In addition, HCV subtype distribution was uneven compared to the general population. Finally, the study findings could contribute to the planning and implementation of health strategies in order to reduce the transmission of infections in the dialysis setting, as well as to monitor and evaluate their effectiveness.

## Acknowledgements

MJP, DF, ER, and FAD are members of the National Research Council (CONICET) Research Career Program. We would like to thank to Mrs. Silvina Heisecke for providing language assistance.

## Declarations

### Availability of data and material

Not applicable

### Code availability

Not applicable

### Authors Contributions

Matías J. PERESON: Analysis and interpretation of data, Drafting the article. Final approval of the version to be published

Alfredo P. MARTÍNEZ: Analysis and interpretation of data, Providing intellectual content of critical importance to the work described. Revising the article. Final approval of the version to be published.

Katia ISAAC: Analysis and interpretation of data, Providing intellectual content of critical importance to the work described. Revising the article. Final approval of the version to be published

Gustavo LAHAM: Analysis and interpretation of data, Providing intellectual content of critical importance to the work described. Revising the article. Final approval of the version to be published.

Ezequiel RIDRUEJO: Analysis and interpretation of data, Providing intellectual content of critical importance to the work described. Revising the article. Final approval of the version to be published.

Gabriel H. GARCIA: Analysis and interpretation of data, Providing intellectual content of critical importance to the work described. Revising the article. Final approval of the version to be published

Diego M. FLICHMAN: analysis and interpretation of data, Providing intellectual content of critical importance to the work described Drafting the article. Final approval of the version to be published

Federico A. DI LELLO: Conception, design, analysis and interpretation of data. Drafting the article. Final approval of the version to be published

## References

1. Marinovich S, Lavorato C, Bisigniano L, Hansen D, Celia E, Tagliafichi V, Rosa Diez G, Fayad A, Haber V. Registro Argentino de Diálisis Crónica SAN-INCUCAI 2017. 2018. https://www.renal.org.ar/Reg_Dial/2017-18/01-2018.pdf. Accessed 4 Jan 2020.

2. Li PK and Chow KM. Infectious complications in dialysis — epidemiology and outcomes. Nat Rev Nephrol. 2011; 8: 77–88.

3. Jefferies M, Rauff B, Rashid H, Lam T and Rafiqet S. Update on global epidemiology of viral hepatitis and preventive strategies. World J. Clin. Cases 2018; 6: 589–99.

4. Boyle S, Lee D and Wyatt C. HIV in the dialysis population: Current issues and future directions. Semin. Dial. 2017; 30: 430–37.

5. Ashkani-Esfahani S, Alavian S and Salehi-Marzijarani M. Prevalence of hepatitis C virus infection among hemodialysis patients in the Middle-East: A systematic review and meta-analysis. World J. Gastroenterol. 2017; 23: 151–66.

6. Jadoul M, Bieber B, Martin P, Akiba T, Nwankwo C, Arduino JM, Goodkin DA, Pisoni RL. Prevalence, incidence, and risk factors for hepatitis C virus infection in hemodialysis patients. Kidney Int. 2019; 95: 939–47.

7. Singh K, Crane M, Audsley J and Lewin S. HIV-Hepatitis B virus co-infection: epidemiology, pathogenesis and treatment. AIDS 2017; 31: 2035–52.

8. Chew KW and Bhattacharya D. Virologic and Immunologic Aspects of HIV-HCV Coinfection. AIDS 2016; 30: 2395–2404.

9. Ganesan M, Poluektova L, Kharbanda K and Osna NA. Human immunodeficiency virus and hepatotropic viruses comorbidities as the inducers of liver injury progression. World J. Gastroenterol. 2019; 25: 398–410.

10. Flichman D, Blejer J, Livellara B, Re VE, Bartoli S, Bustos JA, Ansola CP, Hidalgo S, Cerda ME, Levin AE, Huenul A, Riboldi V, Treviño EM, Salamone HJ, Nuñez FA, Fernández RJ, Reybaud JF, Campos RH. Prevalence and trends of markers of hepatitis B virus, hepatitis C virus and human Immunodeficiency virus in Argentine blood donors. BMC Infect. Dis. 2014; 14: 218.

11. Valtuille R, Frankel F, Gómez F, Moretto H, Fay F, Rendo P, Lef L, Fernández J. The Role of Transfusion-Transmitted Virus in Patients Undergoing Hemodialysis. J Clin. Gastroenterol. 2002; 34: 86–8.

12. Gomez Gutierrez C, Chávez-Tapia N, Ponciano-Rodríguez G, Ponciano-Rodriguez G, Uribe M and Mendez-Sanchez N. Prevalence of hepatitis C virus infection among patients undergoing hemodialysis in Latin America. Ann. Hepatol. 2015; 14: 807–14.

13. Salvatierra K and Florez H. Análisis del virus de la hepatitis C en pacientes en hemodiálisis. Infectio. 2016; 3: 130–37.

14. Neukam K, Ridruejo E, Perez P, Campos RH, Martínez AP and Di Lello FA. Prevalence of hepatitis C virus infection according to the year of birth: Identification of risk groups. Eur. J. Clin. Microbiol. Infect. Dis. 2018; 37: 247–54.

15. Martínez AP, García G, Ridruejo E, Culasso AC, Pérez PS, Pereson MJ, Neukam K, Flichman D, Di Lello FA. Hepatitis C virus genotype 1 infection: Prevalence of NS5A and NS5B resistance-associated substitutions in naïve patients from Argentina. J. Med. Virol. 2019; 91: 1970–78.

16. Kumar S, Stecher G, Li M, Knyaz C, and Tamura K. MEGA X: Molecular Evolutionary Genetics Analysis across computing platforms. Mol. Biol. Evol. 2018; 35: 1547–49.

17. Ronquist F, Teslenko M, van der Mark P, Ayres DL, Darling A, Höhna S, Larget B, Liu L, Suchard MA, Huelsenbeck JP. MrBayes 3.2: efficient Bayesian phylogenetic inference and model choice across a large model space. Syst. Biol. 2012; 61: 539–42.

18. Edey M, Barraclough K and Johnson DW. Review article: Hepatitis B and dialysis. Nephrology 2010; 15: 137–45.

19. Boletín sobre las hepatitis virales en la Argentina. 2019. http://www.msal.gob.ar/images/stories/bes/graficos/0000001592cnt-2019-10_boletin-hepatitis.pdf. Accessed 4 Jan 2020.

20. Fernandez JL, Valtuille R, Hidalgo A, del Pino N, Lef L and Rendo P. Hepatitis G virus infection in hemodialysis patients and its relationship with hepatitis C virus infection. Am. J. Nephrol. 2000; 20: 380–84.

21. Gaite LA, Marciano S, Galdame OA and Gadano AC. Hepatitis C in Argentina: epidemiology and treatment. Hepat. Med. 2014; 6: 35–43.

22. Marinaki S, Boletis JN, Sakellariou S and Delladetsima IK. Hepatitis C in hemodialysis patients. World J. Hepatol. 2015; 7: 548–58.

23. Kershenobich D, Razavi HA, Sánchez-Avila JF, Bessone F, Coelho HS, Dagher L, Gonçales FL, Quiroz JF, Rodriguez-Perez F, Rosado B, Wallace C, Negro F, Silva M. Trends and projections of hepatitis C virus epidemiology in Latin America. Liver Int. 2011; Suppl. 2: 18–29.

24. Simmonds P. The Origin of Hepatitis C Virus. Curr. Top. Microbiol. Immunol. 2013; 369: 1–15.

25. Ruta S and Cernescu C. Injecting drug use: A vector for the introduction of new hepatitis C virus genotypes. World J. Gastroenterol. 2015; 21: 10811–23.

26. Culasso AC, Farías A, Di Lello FA, Golemba MD, Ré V, Barbini L, Campos R. Spreading of hepatitis C virus subtypes 1a and 1b through the central region of Argentina. Infect. Genet. Evol. 2014; 26: 32–40.

27. Di Lello FA, Farias A, Culasso AC, Pérez PS, Pisano MB, Contigiani MS, Campos RH, Ré VE. Changing epidemiology of HCV genotypes in region central of Argentina. Arch. Virol. 2015; 160: 909–15.

28. Boletín sobre el VIH, sida e ITS en la Argentina. 2018. http://www.msal.gob.ar/images/stories/bes/graficos/0000001385cnt-2018-12-20_boletin-epidemiologico-vih-sida-its_n35.pdf. Accesed 4 Jan 2020.

29. Rosenberg AZ, Naicker S, Winkler CA and Kopp JB. HIV-associated nephropathies: epidemiology, pathology, mechanisms and treatment. Nat. Rev. Nephrol. 2015; 11: 150–60.

30. Diana NE and Naicker S. Update on current management of chronic kidney disease in patients with HIV infection. Int. J. Nephrol. Renovasc. Dis. 2016; 9: 223–34.

